# The role of synovial T-cell infiltration following knee joint injury in symptoms and progression to osteoarthritis

**DOI:** 10.1101/19013227

**Authors:** Babak Moradi, Miriam T Jackson, Cindy C. Shu, Susan M Smith, Margaret M Smith, Sanaa Zaki, Hadrian Platzer, Nils Rosshirt, David Giangreco, Carla R Scanzello, Christopher B Little

**Author notes:** **Address for correspondence and reprints: Christopher Little BVMS, PhD**, Raymond Purves Bone and Joint Research Laboratories, Kolling Institute of Medical Research, University of Sydney at Royal North Shore Hospital, St. Leonards, NSW 2065 Australia, **Babak Moradi, M.D.**, University Clinic of Heidelberg, Department of Orthopedics and Trauma Surgery, Schlierbarcher Landstrasse 200a, 69118 Heidelberg, Germany. These authors contributed equally to this research (Babak Moradi, Miriam T Jackson).

## Abstract

**Objectives:** Identification of osteoarthritis(OA)-specific synovial inflammatory pathways, and when in the clinical course they are active, is critical for their utility as therapeutic targets. We directly compared the mononuclear inflammatory/immune-cell responses following joint injury that does and does-not lead to OA, to define bona-fide OA-associated cellular events.

**Methods:** We undertook detailed temporal flow-cytometric and mRNA expression analysis in mice after sham or medial-meniscal-destiblization (DMM) surgery. We compared this with patients with meniscal injury and OA, and evaluated the role of synovial monocytes/macrophages versus lymphocytes in catabolic metalloproteinase secrection *in vitro*. We determined the effect of transient acute or delayed systemic T-cell depletion on DMM-induced OA pathology.

**Results:** OA-inducing/DMM and non-OA-inducing/Sham surgery had identical synovial monocyte/macrophage number, activation and polarization. The number and activation of synovial (not splenic or peripheral-blood) CD4 and CD8 lymphocytes was increased from 1-day after DMM versus Sham, and showed a persistent cyclical elevation throughout OA onset and progression. There was a temporal imbalance in synovial Th17/Treg and Th1/Th2 lymphocytes during DMM-induced OA initiation and progression. We confirmed early post-injury and late-OA CD3/CD8 T-cell responses in synovial tissues from patients, identified an association between CD8 and early post-injury symptoms, and defined a significant role for CD3^+^T-cells in synovial metalloproteinase secretion. Anti-CD3 cell-depletion studies in mice confirmed a key role for the earliest post-injury T-cell response in long-term OA pathology.

**Conclusions:** We identify a hitherto unappreciated pathophysiological role of acute T-cell activation after joint injury in long-term post-traumatic OA risk, providing a novel diagnostic and therapeutic target.

**Key Messages:** *What is already known about this subject?:* The presence of synovitis/joint-inflammation increases the risk not only of osteoarthritis (OA) progression but incident disease. While numerous inflammatory effectors including macrophages and lymphocytes have been identified in OA, their disease-specificity, temporal regulation, and association with risk of pathology onset and progression is lacking.

*How does this study add?:* By directly comparing the mononuclear inflammatory/immune-cell responses following significant joint injury that does (medial-meniscal-destabilization; DMM) and does-not (Sham-surgery) lead to OA in mice, we have defined bona-fide OA-associated cellular events. There was no difference in synovial or systemic monocyte/macrophage cell number, activation or polarization between DMM and Sham, both showing a successful wound-healing response. In contrast, increases in number and activation of synovial Th1- and Th17-CD4, and CD8 T-cells in DMM compared with Sham occurred within the first 3 days, and while recurring cyclically through subsequent disease onset, depletion studies indicated this initial influx was key to long-term ptOA risk.

*How might this impact on clinical practice of future developments?:* Acute increases in synovial T-cells following jont injury may be both a novel marker of OA risk, and a target to reduce long term structural damage.

## Introduction

Osteoarthritis (OA) affects 12-15% of the population causing enormous individual and socioeconomic burden, with no treatments to halt disease onset or progression.[1, 2] Studies in patients and pre-clinical models have implicated OA-associated synovitis as a potential disease-modifying target.[3-8] Synovial inflammatory mediators are more elevated in early OA,[9-11] and patients with synovitis/joint-effusion have faster progression, and more incident OA.[12-14] That inflammation impacts disease initiation is supported by animal studies where synovitis precedes structural hip OA in at-risk dogs,[15] and modulation of specific inflammatory molecules/pathways reduces post-traumatic (pt)OA onset as well as progression in mice.[16] Better understanding of OA-specific inflammation may lead to much needed disease-modifying therapeutics.

Numerous inflammatory effectors have been identified in OA,[3-7] but detailed knowledge of their temporal regulation and association with pathology is lacking, particularly for innate and adaptive immune-cellular responses. Macrophages[17, 18] and lymphocytes[19-22] have been implicated in structural and symptomatic disease as they are present in OA-patient synovium. However, restricted access to preclinical patient samples means the inflammatory-cell events occurring during disease initiation are poorly defined. Occurring after known injury, ptOA offers the potential to study the earliest phases of synovial inflammation,[23] but comparison with “non-OA-inducing” joint injury has not been possible in clinical studies.[24-28]

Surgical destabilization of the medial meniscus (DMM) in mice is a widely used pre-clinical model for investigating ptOA pathobiology and treatment.[29] A key role for specific inflammatory pathways in initiation and progression of DMM-induced ptOA has been demonstrated using genetically-modified mice.[16] We now define a distinct temporal pattern of inflammatory cell influx during initiation and progression of DMM-induced ptOA, that was reflected in meniscal-injury patients, and could be targeted acutely post-injury to modify ptOA risk.

## Methods

### DMM model

Studies were approved by the institutional ethics committee (protocol 1106-012A). Male 10-11-week-old, C57BL6 mice were sourced, housed, and DMM-surgery performed in one knee and Sham-surgery in the other as described.[30, 31] Mice received no post-operative medication, had unrestricted cage-activity, and were sacrificed at day-1, -3, -7, -14, -21, -28, -35, -42, -49, -56, -72, -84, -98 and -112 post-surgery (n=16-22/time). At selected times, age- and sex-matched naïve mice were sacrificed as non-operated-controls (NOC; n=16-22/time). Additional mice had bilateral Sham- or DMM-surgery and were sacrificed at day-14, -42, -84 and -112 post-surgery (n=16/time/group) to evaluate mononuclear cell (MC) changes in peripheral blood (PBMC) and spleen (SpMC). Another cohort of mice (n=20/treatment/time) received a single intra-peritoneal injection of 20µg anti-CD3 (145-2C11; Biolegend) or isotype-control (Armenian-Hamster IgG; Biolegend) 1-week before or 3-weeks after unilateral-DMM, and were sacrificed at day-14, -28, -56, and -112 post-surgery. Eight mice/treatment-group had tactile allodynia measured before and at specified times post-DMM.[32]

### Joint sample preparation

Immediately post-sacrifice synovial tissue (ST: infra-patellar fat-pad + joint capsule + synovial lining) was micro-dissected en-bloc.[30] ST from 6 NOC, Sham and DMM joints at day-7, -14, -35, -49 and - 112 were used for qRT-PCR [30, 31] to quantify expression of *Cd3, Cd4, Cd8, Il1, Il6*, and *Tnf* relative to *Gapdh* (mouse-specific primers; Supplementary Table I). ST from 16 animals/group were used for flow-cytometry (4 “biological-replicates”/timepoint consisting of pooled ST from 4 mice). After ST removal, remaining joint tissues were processed and sagittal sections every 80µm through the medial femoro-tibial joint stained with toluidine-blue/fast-green for histopathology scoring.[30, 31]

### Flow-cytometry

PBMCs, SpMCs, and STMCs were isolated, live cells counted, and subtypes quantified using cell surface markers (Figure 2&3).[30] A total of 10^5^ events were collected (four-colour flow-cytometer; Becton-Dickinson), and population analysis performed (FlowJo, Treestar). Viable (7-aminoactinomycin-D-negative) CD45^+^ MCs were gated on forward/side scatter as monocytic (CD11b^+^) or lymphocytic (CD3^+^) then: CD11b^+^F4/80^-^Ly6c^+^ resident-monocytes, CD11b^+^F4/80^+^Ly6c^high^ inflammatory-monocytes, CD11b^+^F4/80^+^Ly6c^low^ activated-macrophages (M1(F4/80^+^CD11c^+^) or M2(F4/80^+^CD206^+^)), CD3^+^CD4^+^CD8^-^ T-helper, and CD3^+^CD4^-^CD8^+^ cytotoxic-T-cells. For intracellular staining, cells were stimulated (5h, 50ng/ml phorbol-myristate-acetate, 1μg/ml ionomycin, BD-Biosciences) before addition of brefeldin-A (5μg/ml, 5h; Sigma-Aldrich). Following surface-marker staining, cells were fixed, permeabilized and stained with BV-421-labelled-anti-IFN-γ (“Th1”), APC-labelled-anti-IL-4 (“Th2”), PE-labelled-anti-IL-17A (“Th17”) or Alexa-488-labelled-anti-FoxP3 (“Treg”).

**Figure 1:**
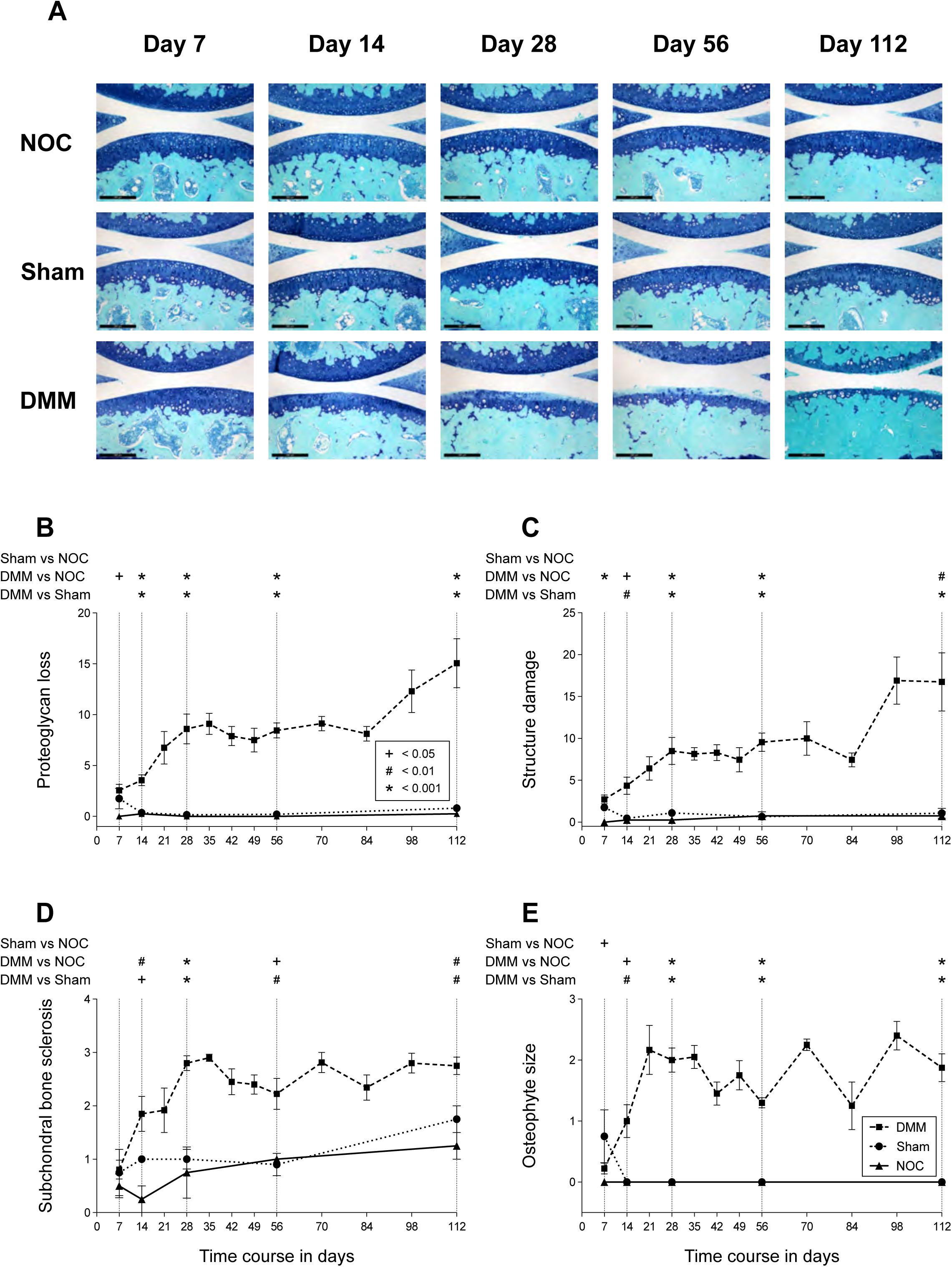
Progressive osteoarthritis structural pathology occurs after destabilization of the medial meniscus (DMM) but not Sham surgery. **(A)** Representative images of toluidine blue/fast-green stained sections of medial femorotibial joints of mice 7-112 days after Sham or DMM surgery compared with age-matched non-operated controls (NOC; scalebar = 200µ)). Progressive loss of cartilage proteoglycan staining followed by non-calcified cartilage erosion only occurs in DMM and is accompanied by increased subchondral bone sclerosis and loss of epiphyseal marrow. Two observers blinded to injury and time scored the tibial **(B)** cartilage proteoglycan loss, **(C)** cartilage structural damage, **(D)** subchondral bone sclerosis, and (**E**) osteophyte size. Data is shown as the mean ± SEM; NOC = triangle and solid line, Sham = circle and dotted line, DMM = square and dashed line. Statistical comparison was performed for those time points where scores for all three groups were available (highlighted with a vertical dashed line); n = 10 for DMM and Sham, and n = 4 for NOC; *=*P*≤0.05; +=*P*≤0.01; #=*P*≤0.001.

**Figure 2:**
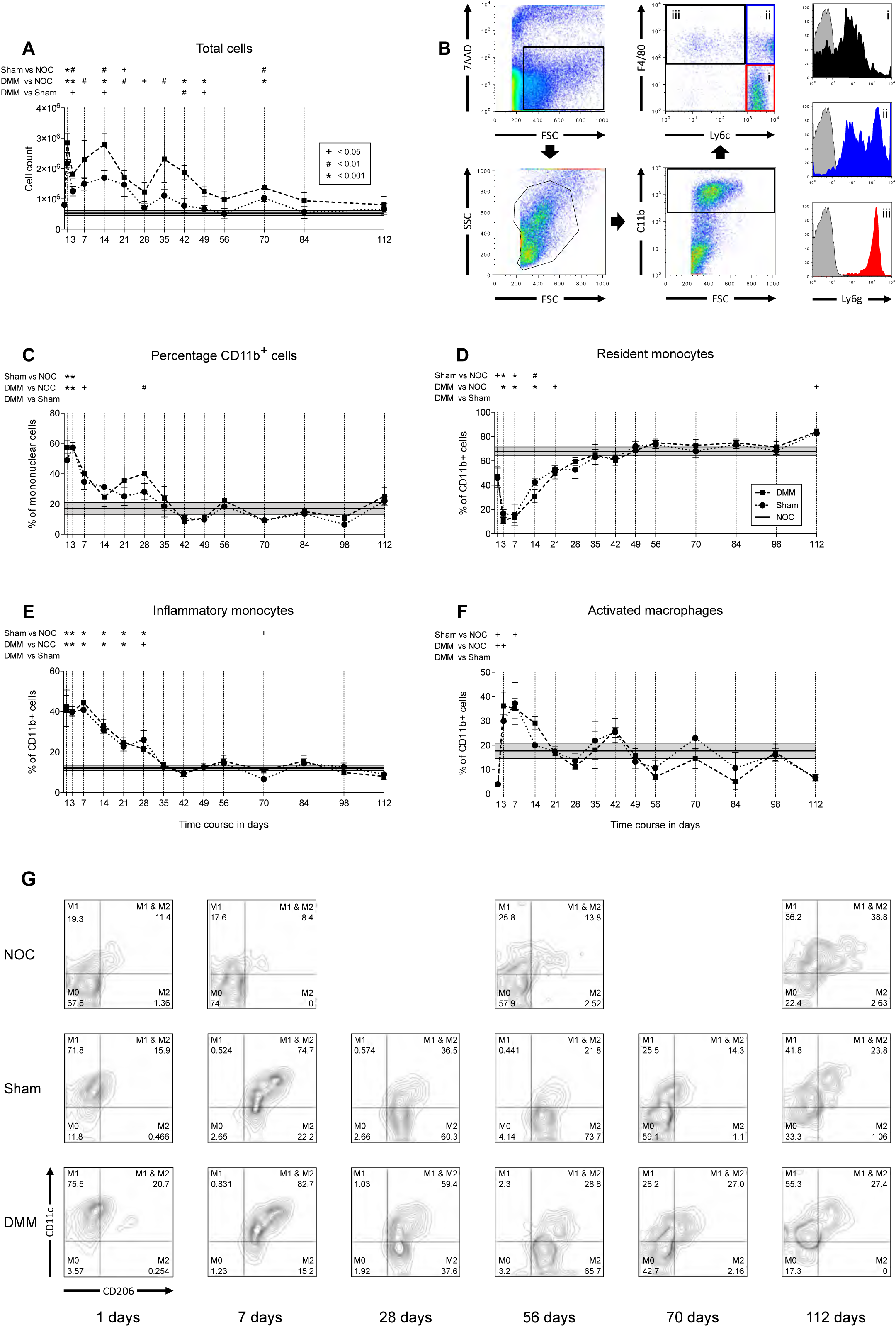
DMM results in greater cyclical synovial mononuclear cell influx than Sham surgery but no difference in monocyte/macrophage activation or polarization. **(A)** Both Sham and DMM surgery resulted in cyclical increases in mononuclear cell number in the synovial tissue compared with non-operated controls (NOC), however the effect was more pronounced and persistent with DMM. There was no difference in synovial cell number in NOC over time so a mean value (solid line ± SEM represented by grey shading) was calculated and used for comparison with Sham (circle with dotted line) and DMM (square with dashed line) at each time point indicated with a vertical line (n = 4 [each a pool of 4 mice]/time/surgery; *=*P*≤0.05; +=*P*≤0.01; #=*P*≤0.001). **(B)** Isolated synovial cells were gated for flow cytometric analysis using the forward scatter profile and 7-AAD staining to exclude debris and dead cells. Monocyte/macrophage cell populations were then defined by cell surface markers (cut offs established using isotype control antibodies; bold arrows indicate the sequential gating strategy): (i) resident monocytes (CD11b^+^F4/80^-^Ly6c^+^), (ii) inflammatory monocytes (CD11b^+^F4/80^+^Ly6c^high^), (iii) activated macrophages (CD11b^+^F4/80^+^Ly6c^low^), which were further distinguished by their relative expression of Ly6g (right panel; colours according to gate i, ii, or iii). The frequency of synovial CD11b^+^ cells **(C)**, resident monocytes (**D)**, inflammatory monocytes **(E)**, and activated macrophages **(F)**, showed distinct temporal changes after surgery compared with NOC but with no significant difference between Sham and DMM (n and analyses as described for panel A). (**G)** Activated macrophages (CD11b^+^F4/80^+^Ly6c^low^) in Sham and DMM showed a very similar temporal pattern of polarization from M0 (CD11c^-^/CD206^-^) to M1 (CD11c^+^/CD206^-^), M1/2 (CD11c^+^/CD206^+^) and finally M2 (CD11c^-^/CD206^+^).

**Figure 3:**
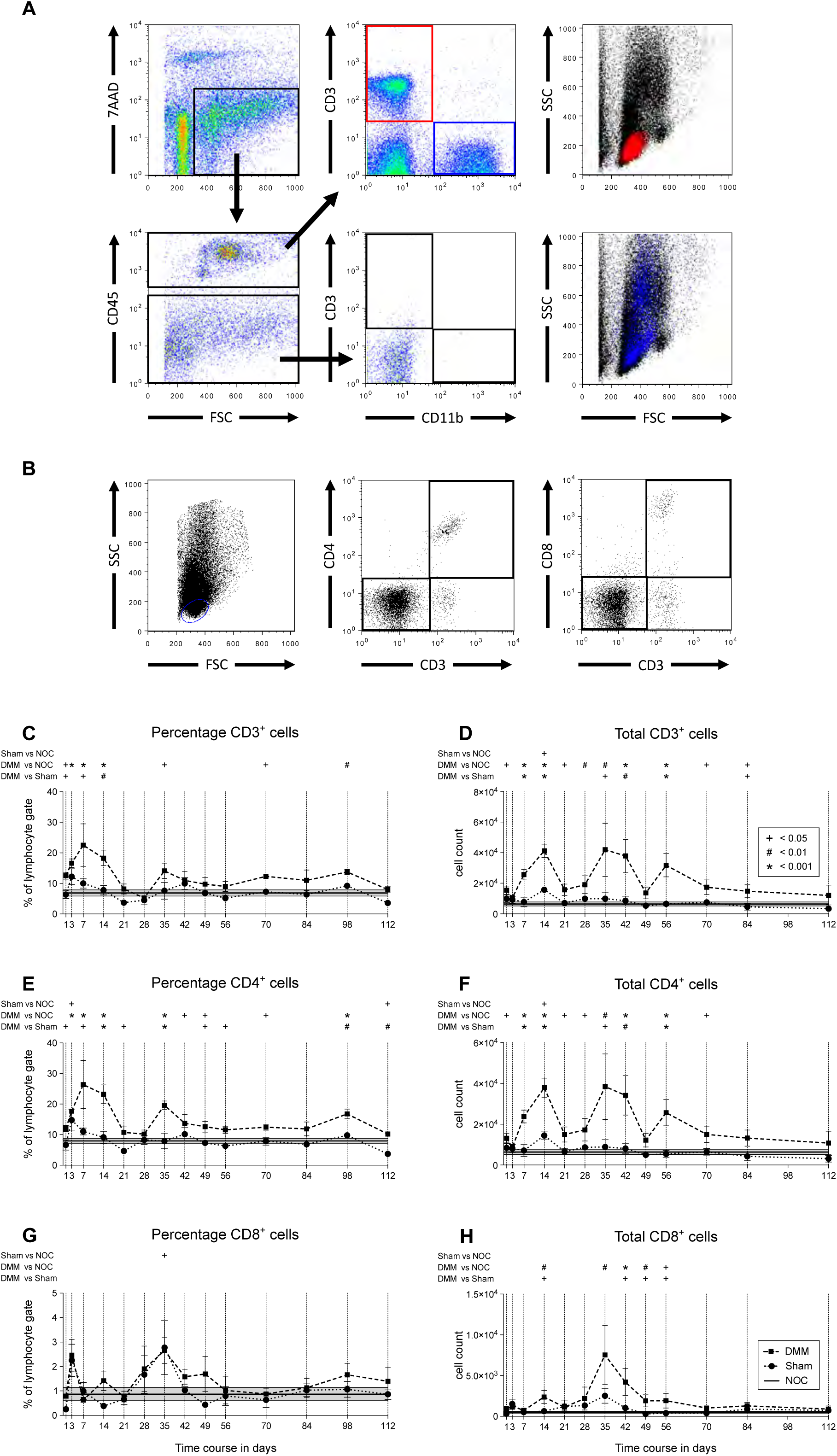
DMM results in significantly greater cyclical synovial T-cell cell influx than Sham surgery. **(A)** Viable mononuclear cells were gated based on the negative 7-AAD staining and further through the CD45 marker. CD3 and CD11b were used as broad markers for lymphocytes and monocytes/macrophages cells, respectively. These two distinct cell populations showed no overlap and were all CD45^+^, with little or no CD3 or CD11b staining in the CD45^-^ population. The right column shows the cell size and granularity of CD3^+^ (red) and CD11b^+^ (blue) cells. Cells in the “lymphocyte gate” based on forward and side scatter were further defined as CD3^+^CD4^+^CD8^-^ T-helper cells and CD3^+^CD4^-^CD8^+^ cytotoxic T-cells **(B)**. The frequency **(C, E, G)** and absolute number **(D, F, H)** of synovial CD3^+^ lymphocytes **(C, D)**, CD4+ T-helper **(E, F)** and CD8+ cytotoxic T-cells **(G, H)**, showed distinct temporal changes after surgery compared with NOC, with significantly greater numbers at multiple timepoints in DMM compared with Sham. There was no difference in synovial lymphocytes in NOC over time so a mean value (solid line ± SEM represented by grey shading) was calculated and used for comparison with Sham (circle with dotted line) and DMM (square with dashed line) at each time point indicated with a vertical line (n = 4 [each a pool of 4 mice]/time/surgery; *=*P*≤ 0.05; +=*P*≤0.01; #=*P*≤0.001).γ

### Human samples

Suprapatellar ST collected in the course of previous studies from three patient cohorts (Supplementary information) were analysed: 16 “*advanced knee OA”*,[24] 19 “*meniscal-tear early-OA”*,[24, 33] and 13 “*meniscal-tear no-OA”*.[26, 27] Aliquots of RNA from de-identified patients were provided with clinical data (Supplementary Table II), and *CD3, CD4* and *CD8* quantified by qRT-PCR (human-specific primers; Supplementary Table I).

ST from 12 OA patients undergoing uni-compartmental or total knee replacement (University Hospital Heidelberg; ethics #S-156/2014; Supplementary Table III), were digested (collagenase B [Roche], 6.7µg/mg 37°C 2hr), and MCs isolated by centrifugation. Magnetic-activated-cell-sorting depletion of CD45^+^, CD14^+^ or CD3^+^ cells was done according to the manufacturer’s instructions (Miltenyi Biotec). Native and depleted samples were cultured (24 hours, RPMI/10%-FCS/1%-Penicillin-Streptomycin; 10^6^ cells/ml), and the concentration of MMP-1 (Sigma-RAB0361-1KT), MMP-3 (Sigma-RAB0367-1KT), MMP-9 (Sigma-RAB0372-1KT), and ADAMTS-5 (LOXO-6SEK205HU) in conditioned media quantified by ELISA.

### Statictical Analysis

The minimum number of animals and/or biological replicates to detect differences with >80% power (two-sided, alpha 0.05) was determined *a priori* using previous data:[30, 31, 34] n = 5/group for histopathology, n = 4/group for FACS, n = 6/group for gene expression, and n = 8/group for mechanical allodynia. Animals were randomly assigned to groups, and investigators were blinded to these during data acquisition. Data from all animals and human subjects was used for analysis (no outlier removal), with the number of replicates provided in methods, figure legends and tables. Following QQ plots and Shapiro-Wilk tests, normally distributed data (flow-cytometry, ELISA) was analysed by ANOVA with Bonferroni and Tukey correction, while non-parametric outcomes (histopathology, gene-expression) were evaluated using Friedman tests (Prism, GraphPad). Spearman correlations between clinical outcomes and mRNA expression were calculated using SPSS (IBM Inc). For all analyses p≤0.05 was considered statistically significant.

## Results

### OA histopathology

Progressively worsening medial femoro-tibial OA pathology including signficanty greater cartilage proteoglycan loss and structural damage, subchondral bone sclerosis and osteophyte formation was seen in DMM versus NOC (from day-7) and Sham (from day-14) (Figure 1).

### Synovial mononuclear cell number

The number of STMCs in NOC did not change over time (*P*=0.84). STMCs increased markedly 1-day following Sham and DMM, with cyclical decreases (day-3, -28, -56 and -84) and increases (day-14, -35 and -70) thereafter (Figure 2A). Resolution phases were greater and increases less in Sham, such that numbers returned to NOC levels after day-21, while remaining elevated in DMM versus NOC (day-1-70), and Sham (day-3-49).

### Monocytes/macrophages

Frequency of systemic CD11b^+^ monocytes/macrophages were similar in NOC, Sham and DMM: 41.6±27.1%, 35±17.5% and 30.6±12.9% of PBMC, respectively; 6.9±0.6%, 9.8±1.5%, and 5.8±0.6% of SpMC, respectively; no temporal differences within any group. This also applied to systemic CD11b^+^ subtypes: Ly6c^+^resident-monocytes (NOC:43.7±11%, Sham:49±13%, DMM:44±10%), F4/80^+^Ly6c^high^inflammatory-monocytes (NOC:26.6±3%, Sham:30.4±6%, DMM:29.1±3.5%), and F4/80^+^Ly6c^low^activated-macrophages (NOC:19.8±7.8%, Sham:17.5±6.8%, DMM:23.1±6.3%). In contrast, CD11b^+^ STMCs increased immediately post-surgery followed by diminishing cyclical peaks (day-3, -28, -56, and -112), but with no difference between Sham and DMM (Figure 2C). Resident-monocytes made up 60-80% of CD11b^+^ STMCs in NOC at all times, but decreased acutely in Sham and DMM (<20% by day-3) before returning to NOC levels (day-28-35; Figure 2D). Inflammatory-monocytes were elevated immediately post-surgery before returning to NOC levels at day-35 (Figure 2E). Activated-macrophages peaked 3-7 days post-surgery before returning to NOC levels at day-21 (Figure 2F). There was no difference between Sham and DMM in any subtype at any time.

Polarization of ST-macrophages into M1 and M2 subtypes showed distinct temporal patterns in NOC versus DMM and Sham, but no diference between surgeries (Figure 2G). In NOC the majority of ST-macrophages expressed neither marker up to day-56 (M0:57-74%), with 17-26% M1, 8-14% positive for both (M1/2), and only 0.1-2.5% M2. With ageing (day-112) increases in M1 (36%) and M1/2 (39%) occurred in NOCs. In both DMM and Sham there was a marked increase in M1 at day-1 (72-76%), followed by a temporal transition to M1/2 (75-83% at day-7), M2 (66-73% by day-56), and thereafter to day-112 NOC levels.

### Lymphocytes

There was a cyclic increase in ST-lymphocytes after surgery (Figure 3): peaks at day-7, -35, and -98. The percentage of ST-CD3^+^-lymphocytes did not differ between Sham and NOC at any time but was greater in DMM than NOC (day-1-98), and Sham (day-1-14; Figure 3C). As a percentage of the CD3^+^-lymphocytes, CD4^+^-T-cells were increased in DMM compared with NOC and Sham from day-1-112 (Figure 3E). While showing similar cyclic fluctuation, the percentage of CD8^+^-T-cells did not differ between DMM and Sham (Figure 3G). However, CD3^+^, CD4^+^ and CD8^+^ T-cell numbers (rather than %) were greater in DMM versus Sham from day-1-84 (Figure 3D, F, H).

Splenic and ST T-helper cells were further categorized post-DMM as Th1(IFN^+^), Th2(IL-4^+^), Th17(IL-17^+^) or Treg(FoxP3^+^) (Figure 4A). The percentage of all subtypes was almost always lower in the spleen, and when this was not the case anti-inflammatory cell-types predominated (Table I). There was little change in splenic CD4^+^ subtypes over time (all <5%; Figure 4B), and pro-inflammatory imbalances (Th1/Th2, Th17/Treg) were generally <2-fold (Supplemantary Table IV). In the ST however, large temporally-distinct changes in T-helper subtypes occurred (Figure 4B, Table I). Th17 cells predominated at day-7 post-DMM (19%) with levels remaining >10% thereafter, while Tregs were least common at day-7 (1.4%) increasing to 25% by day-35 then decreasing to <5% by day 84-112. Th1 showed the most dramatic post-DMM increase reaching >70% of CD4^+^ T-cells at day-70 before returning to ∼20% at day-112, and while the pattern was broadly similar Th2 changes were smaller (peaking at ∼33% at day-70). Unlike the spleen there was a marked pro-inflammatory imbalance in ST-CD4^+^-T-cell subtypes post-DMM: Th1 significantly >Th2 at day-35, -56, -70, and - 98; Th17 significantly >Treg at day-1, -70, -84 and -98 (Figure 4B; Supplementary Table IV).

**Table I:** Comparison of the % of different CD4 T-cell subtypes (Th1(IFN-γ^+^), Th2(IL-4^+^), Th17(IL- 17^+^), Treg(FoxP3^+^)) in the spleen versus synovium in mice at different times (day 7, 35, 56, 70, 84, 98 and 112) post-DMM surgery. Data shows the mean ± standard deviation (SD (range)) of the % of each cell-type; n = 4-8 for each tissue at each time.

| Cell-type and time post-DMM | Spleen | Synovium | P-Value |
| --- | --- | --- | --- |
| | Mean ± SD (range) | Mean ± SD (range) | (spleen vs synovium) |
| Th1(IFN-$\gamma^+$) day-7 | 2.3 ± 0.3 (2.0 – 2.7) | 13.9 ± 3.6 (9.0 – 18.6) | **<0.001** |
| Th1(IFN-$\gamma^+$) day-35 | 2.9 ± 2.0 (1.7 – 5.8) | 67.7 ± 7.9 (60.8 – 79.0) | **<0.001** |
| Th1(IFN-$\gamma^+$) day-56 | 1.1 ± 1.2 (0.4 – 2.8) | 39.0 ± 16.0 (19.1 – 58.3) | **0.003** |
| Th1(IFN-$\gamma^+$) day-70 | 2.1 ± 0.4 (1.5 – 2.5) | 71.3 ± 11.4 (57.6 – 83.3) | **<0.001** |
| Th1(IFN-$\gamma^+$) day-84 | 3.3 ± 0.9 (2.3 – 4.4) | 31.8 ± 17.9 (15.0 – 60.4) | **0.003** |
| Th1(IFN-$\gamma^+$) day-98 | 0.7 ± 0.4 (0.1 – 1.2) | 7.9 ± 1.9 (5.1 – 9.9) | **<0.001** |
| Th1(IFN-$\gamma^+$) day-112 | 2.1 ± 0.9 (0.8 – 3.9) | 20.7 ± 14.0 (7.5 – 38.4) | 0.076 |
| Th2(IL-4$^+$) day-7 | 2.8 ± 0.4 (2.3 – 3.5) | 11.9 ± 9.1 (4.3 – 26.3) | **0.025** |
| Th2(IL-4$^+$) day-35 | 2.1 ± 1.5 (1.1 – 4.4) | 20.4 ± 5.0 (16.1 – 27.5) | **<0.001** |
| Th2(IL-4$^+$) day-56 | 4.5 ± 1.2 (3.5 – 6.2) | 0.9 ± 0.4 (0.3 – 1.3) | **0.001** |
| Th2(IL-4$^+$) day-70 | 1.1 ± 0.2 (0.9 – 1.3) | 32.9 ± 7.9 (24.1 – 40.2) | **0.004** |
| Th2(IL-4$^+$) day-84 | 2.2 ± 0.7 (1.2 – 3.5) | 23.1 ± 5.0 (18.3 – 28.6) | **<0.001** |
| Th2(IL-4$^+$) day-98 | 1.5 ± 0.8 (0.6 – 2.8) | 2.4 ± 0.4 (2.0 – 3.0) | **0.023** |
| Th2(IL-4$^+$) day-112 | 1.5 ± 0.9 (0.5 – 3.6) | 2.5 ± 1.2 (1.3 – 4.0) | 0.088 |
| Th17(IL-17$^+$) day-7 | 1.4 ± 0.3 (1.0 – 1.9) | 19.1 ± 8.1 (12.7 – 32.3) | **<0.001** |
| Th17(IL-17$^+$) day-35 | 4.1 ± 1.1 (2.8 – 5.5) | 20.0 ± 4.8 (13.7 – 24.5) | **0.005** |
| Th17(IL-17$^+$) day-56 | 0.6 ± 0.5 (0.0 – 1.1) | 10.0 ± 3.1 (6.3 – 13.4) | **0.008** |
| Th17(IL-17$^+$) day-70 | 3.8 ± 1.7 (2.7 – 6.3) | 22.4 ± 7.1 (14.2 – 31.5) | **0.011** |
| Th17(IL-17$^+$) day-84 | 1.2 ± 0.5 (0.5 – 2.1) | 5.6 ± 3.4 (2.7 – 10.5) | **0.009** |
| Th17(IL-17$^+$) day-98 | 2.6 ± 0.9 (1.0 – 3.6) | 12.7 ± 1.9 (9.8 – 14.5) | **<0.001** |
| Th17(IL-17$^+$) day-112 | 3.6 ± 1.5 (1.6 – 6.2) | 9.6 ± 4.3 (3.3 – 12.3) | 0.062 |
| Treg(FoxP3$^+$) day-7 | 1.1 ± 0.6 (0.8 – 2.6) | 1.4 ± 0.6 (0.4 – 2.1) | 0.45 |
| Treg(FoxP3$^+$) day-35 | 2.1 ± 0.3 (1.7 – 2.4) | 25.7 ± 8.0 (19.3 – 37.1) | **0.001** |
| Treg(FoxP3$^+$) day-56 | 1.6 ± 1.1 (0.8 – 3.2) | 14.0 ± 5.7 (8.8 – 21.9) | **0.005** |
| Treg(FoxP3$^+$) day-70 | 1.7 ± 0.2 (1.4 – 1.9) | 11.9 ± 2.5 (8.7 – 14.1) | **0.004** |
| Treg(FoxP3$^+$) day-84 | 0.3 ± 0.1 (0.2 – 0.4) | 0.0016 ± 0.0016 (0.0001 – 0.005) | **<0.001** |
| Treg(FoxP3$^+$) day-98 | 2.3 ± 1.4 (0.9 – 5.1) | 0.7 ± 1.1 (0.0 – 2.4) | **0.021** |
| Treg(FoxP3$^+$) day-112 | 3.1 ± 0.6 (2.2 – 4.4) | 4.3 ± 4.0 (1.0 – 10.0) | 0.61 |

**Figure 4:**
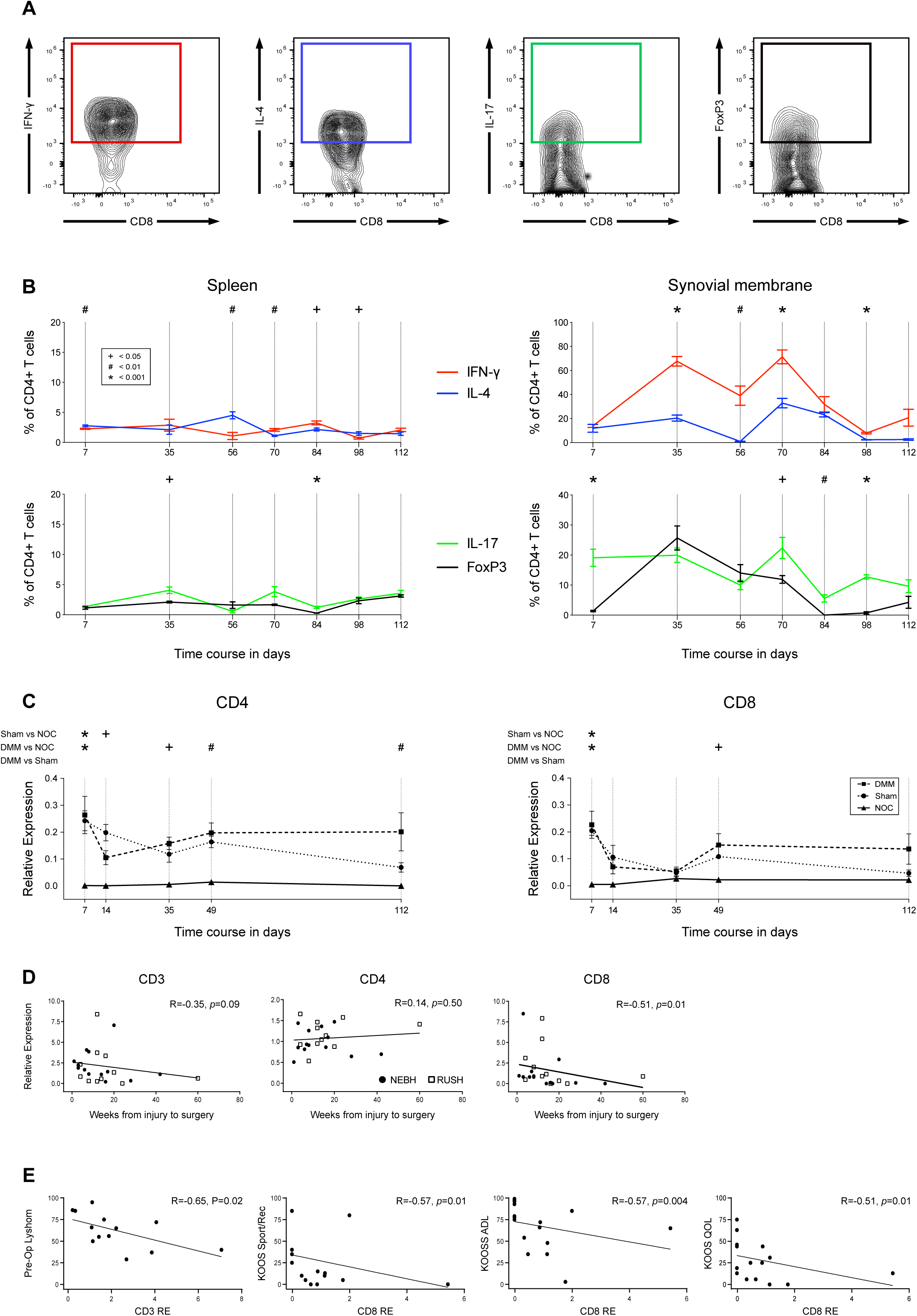
T-cell changes in post-traumatic osteoarthritis (ptOA) are localised to the joint, show temporally distinct Th1/Th2 and Th17/Treg imbalance, and are associated with worse clinical symptoms in patients. **(A)** CD45^+^CD3^+^CD4^+^CD8^-^ T-cells subsets were defined by production of their specific cytokines and transcription factors: Th1-IFNγ^+^, Th2-IL-4^+^, Th17-IL-17a^+^, and Treg-FoxP3+. **(B)** DMM surgery resulted in little temporal change in these CD4^+^ T-cell subsets in the spleen but pro-inflammatory imbalances in Th17/Treg acutely after injury and in late disease, while a Th1/Th2 imbalance predominated during the progression of ptOA structural damage. Data is shown as the mean ± SEM (n = 4 replicates [each a pool of 6 mice]/time/surgery) with differences between Th17:Treg and Th1:Th2 analysed as each time (*=*P*≤0.05; +=P≤0.01; #=*P*≤0.001). **(C)** Analysis of synovial *Cd4* and *Cd8* mRNA expression in mice showed a cyclic pattern (similar to that seen with flow cytometry) with more persistent elevation in DMM than Sham. Data shows mean ± SEM for NOC (triangle with solid line), Sham (circle with dotted line), and DMM (square with dashed line), with comparisons between groups at each time point indicated with a vertical line (n = 6 individual mice/time/surgery; *=*P*≤0.05; +=*P*≤0.01; #=*P*≤0.001). **(D)** Expression of *CD3* and *CD8* (but not *CD4*) in synovial tissue collected at the time of arthroscopic partial meniscectomy in patients without or with early OA (NEBH and Rush cohorts, respectively), showed higher levels with a shorter time after injury. **(E)** Higher *CD3* (NEBH patients) and *CD8* (Rush patients) were significantly correlated with worse pre-operative symptoms (lower Lysholm or KOOS scores).

### ST-lymphocyte-marker expression

Expression of *Cd4* and *Cd8* was markedly increased at day-7 in both Sham and DMM (Figure 4C), decreasing by day 14-35, before increasing again, particularly in DMM, such that by day-112 expression was higher than Sham, reflecting flow-cytometry data (Figure 3). Expression of *CD3, CD4* and *CD8* mRNA was therefore measured in ST from patients after meniscal injury with no (“NEBH”) or early (“Rush”) OA, as well as with advanced-OA. *CD3* and *CD4* were detected in 95-100% of all patients (no difference between groups), while the percentage of *CD8*-positive advanced-OA patients (100%) was greater than meniscal injury (NEBH 77%, Rush 63%). There was no between-group difference in mean *CD3, CD4* or *CD8* expression, however *CD3* and *CD8* were higher with shorter time post-injury (Figure 4D). Lymphocyte marker expression was significantly correlated with pre-operative symptoms: NEBH-patient *CD3* and Lysholm score; Rush-patient *CD8* and KOOS-pain, KOOS-ADL, KOOS-Sport/recreation, and KOOS-QoL scores (Figure 4E; Supplemantary Table V).

### Treating OA by lymphocyte depletion

Magnetic-activated-cell-sorting selectively depleted CD45^+^, CD14^+^ or CD3^+^ cells from human OA-ST (Figure 5A). Depleting all hemopoietic-lineage (CD45^+^) cells or only CD14^+^ monocytes/macrophages, markedly decreased MMP1, MMP3 and MMP9 secretion (Figure 5B). Removing only CD3^+^ lymphocytes had less effect on MMP3 but equivalent decreases in MMP1 and MMP9. In light of this *in vitro* data, we evaluated whether lymphocyte depletion would modify DMM-induced ptOA, and compared targeting the immediate post-injury influx versus that occurring later: pre-DMM versus 3-weeks post-DMM anti-CD3. Treatment at either time depleted splenic CD3^+^ and CD4^+^ T-cells for 2-weeks, and CD8^+^ T-cells for up to 8-weeks (Figure 5C), but there was no significant effect on ST T-cells (Figure 5D). Neither treatment protocol altered subchondral bone remodeling or osteophyte size, but osteophyte maturation from cartilage to bone was delayed: week-2 and week-8 after pre- and post-DMM treatment, respectively (Figure 5E). There was no effect of post-DMM anti-CD3 on cartilage pathology, however, targeting the initial T-cell response reduced 2-week proteoglycan loss compared with IgG, slowed progressive 8-16-week proteoglycan loss (Figure 5F) and cartilage structural damage (Figure 5G), and reduced late-stage (16-week) cartilage damage (Figure 5G).

**Figure 5:**
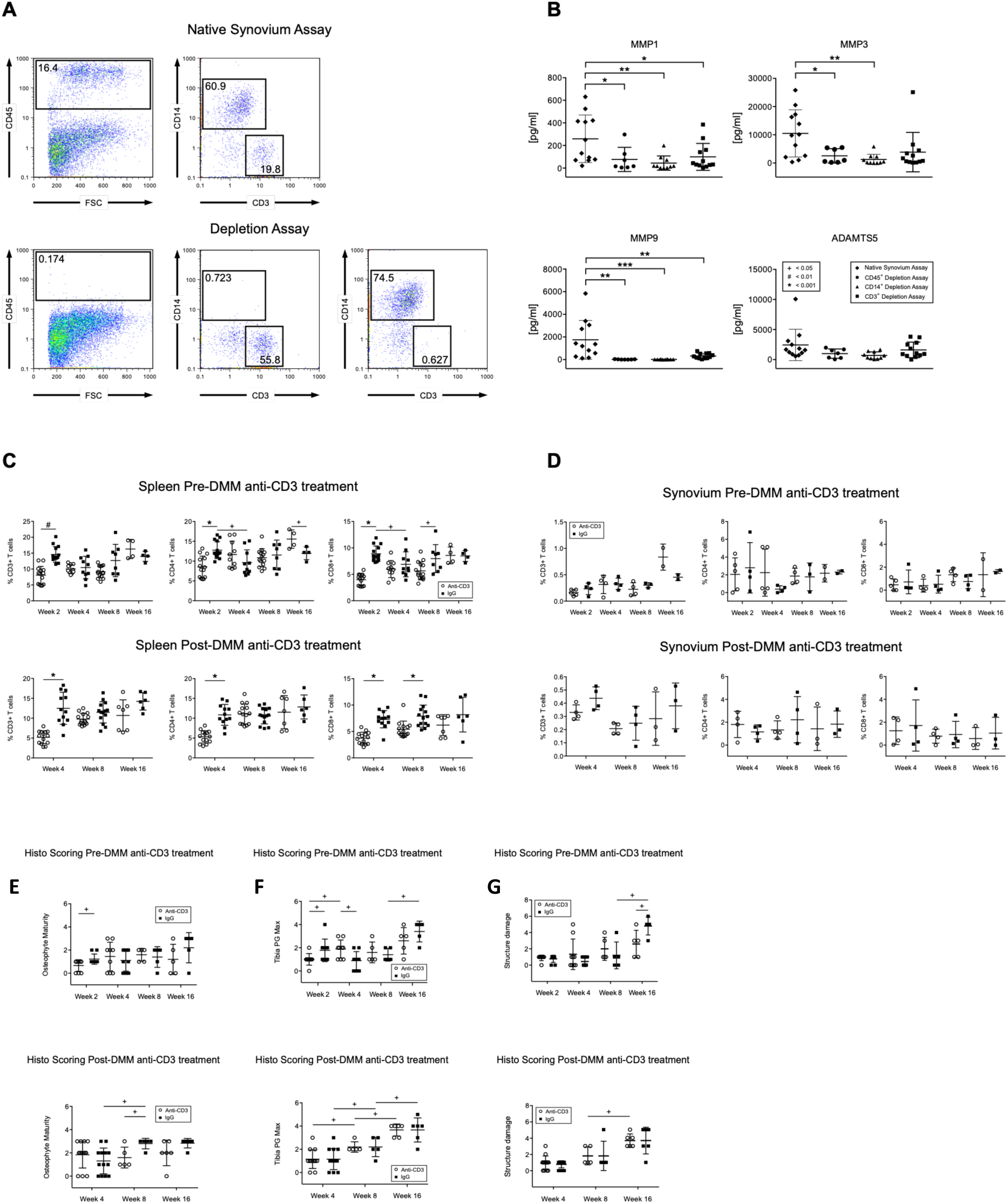
Depletion of T-cells significantly reduces osteoarthritic synovial cell metalloproteinase secretion and late stage progression and severity of DMM-induced post-traumatic OA. **(A)** Selective and efficient depletion of CD45^+^, CD14^+^ or CD3^+^ cells in isolates from human OA synovium was achieved with magnetic-activated-cell-sorting. **(B)** Compared with native synovial tissue cell isolates, significant reduction in MMP1, MMP3 and MMP9 (but not ADAMTS5) secretion into conditioned media occurred with depletion of all haemopoietic cells (CD45^+^), only monocytes/macrophages (CD14^+^), or only lymphocytes (CD3^+^). Data shows all individual patients as well as mean ± SEM (n = 12 group; *=*P*≤0.05, **=*P*≤0.01, ***=*P*≤0.001). **(C)** Systemic anti-CD3 administered either 1 week Pre- or 3 weeks Post-DMM significantly depleted splenic CD3^+^ and CD4^+^ T-cells for 2-weeks post-treatment, and CD8^+^-T-cells for 8- (Pre-DMM treatment) or 5-weeks (Post-DMM treatment) compared with control antibody. With pre-DMM treatment the CD4^+^ T-cells rebounded and were increased compared with control antibody at week-16. Data shows all individual replicates as well as mean ± SEM (n = 4-12 individual mouse spleens/time/treatment; *=*P*≤0.05; +=*P*≤0.01; #=*P*≤0.001). **(D)** Despite effects on splenic T-cell cells, there was no change in CD4^+^ or CD8^+^ T-cells in the synovial tissues following systemic anti-CD3 administered either 1 week Pre- or 3 weeks Post-DMM (n = 2-5 replicates [each a pool of 4 mice]/time/treatment). **(E)** Osteophyte maturation was reduced at 2- and 8-weeks post-DMM by anti-CD3 administered 1 week Pre- or 3 weeks Post-DMM, respectively. **(F)** While cartilage proteoglycan loss was unaffected by anti-CD3 given Post-DMM, it was reduced at 2-weeks when administered Pre-DMM which also slowed progressive loss from 8-16 weeks. **(G)** There was no effect of post-DMM anti-CD3 on tibial cartilage structural pathology, however pre-surgical treatment significantly slowed late stage cartilage structural damage progression from 8-16 weeks, and reduced severity of 16-week cartilage damage compared with IgG control. Histology data (panels **E-G**) shows all individual mice as well as mean ± SEM (n = 5-8 mice/time/treatment; *=*P*≤0.05; +=*P*≤0.01).

## Discussion

Burgeoning evidence implicates synovial inflammation not just in OA symptoms but structural disease pathophysiology.[3-8] Failure of non-specific anti-inflammatory approaches (NSAIDs, corticosteroids) to modify OA pathology, and experience with biologics in rheumatoid arthritis,[35] suggest targeting of particular inflammatory pathways/effectors will be needed for disease-modification. Moreover, the disease-specificity of these pathways and knowledge of when in the clinical course they are active is critical for therapeutic utility. Directly comparing mononuclear inflammatory/immune-cell responses following significant joint injury that does (DMM-surgery) and does not (Sham-surgery) lead to OA, allowed us to define bona-fide OA-associated events.

Despite equivalent surgical joint insult and the same acute histologically-graded inflammation,[30] flow-cytometry revealed a more profound and distinct STMC influx in DMM compared with Sham, not only with later structural disease but within the first 3 days post-injury, well before OA-histopathologic change. The current paradigm posits that inflammation in joint injury follows a classical wound-healing pattern: tissue trauma inciting an innate-immune response involving activation of the complement cascade, with neutrophil and then monocyte/macrophage influx and activation.[5, 11, 23, 36] Non-OA inducing injury would be considered a healing wound where inflammation resolves as injured tissues repair and remodel, while damage-asscociated-molecular-patterns (DAMPS) released from degrading joint tissues after OA-inducing injury cause ongoing innate-immune activation that drives an adaptive immune response, as in chronic/non-healing wounds.[37, 38] However, we found that with OA-inducing injury (DMM-surgery), ST-monocyte/macrophage influx and polarization followed the same “successful” wound-healing pattern as Sham-surgery: acute M1-inflammatory response (peak day 3-7), followed by progressive (4-5 weeks) reparative-M2-polarization, and then return to age-matched naïve levels. In contrast, there was a rapid (day-1) and chronic cyclical elevation of ST-lymphocytes that, unlike the innate-immune response, differentiated OA-inducing/DMM from resolving/Sham injury. This implicates a more important role for the adaptive-immune system in ptOA risk following joint injury, and that this occurs much earlier than previously suspected.[5, 39]

Rather than a unimodal “insult-inflammation-resolution” model,[38] there were cyclical increases and decreases in STMCs after joint injury, that were accentuated in DMM and particularly apparent for lymphocytes. The cyclic pattern could be related to temporal release of DAMPs from different tissues or different DAMPs released from one progressively damaged tissue.[36, 40] The slight lag between monocyte/macrophage and lymphocyte peaks (Figure 2 and 3) is consistent with the former secreting chemokines that attract the latter in a coordinated injury-response.[38] However, despite equivalent monocyte/macrophage number, activation/polarization and associated *Il1* and *Il6* expression (Supplementary Figure 1A) in the immediate post-injury period, there was significantly greater acute ST-lymphocyte influx in DMM versus Sham. This could be associated with specific DAMP/antigen release from tissues only injured in DMM (menisco-tibial ligament, destabilized meniscus[41]) and/or chemokines being secreted from mechanically injured chondrocytes which occurs before histomorphologic pathology.[42]

Analysis of PBMCs and SpMCs indicated a local rather than systemic response to DMM even with bilateral knee injury, and that OA-associated adaptive-immune response in one joint does not lead to widespread change, unlike rheumatoid arthritis.[43] Our data contrasts with spontaneous-OA in the STR/ort mouse, where macrophage and T-cell ST-infiltration is accompanied or preceded-by systemic increases.[44, 45] Joint pathology in the STR/ort mouse is driven at least in part by hyperlipidaemia and associated hypercholesteremia, hyperinsulinemia, insulin-resistance and systemic inflammation.[45-47] This “metabolic-like-syndrome” is a significant risk for OA in patients,[48] and absence of such co-morbidities in the non-obese C57BL6 mice in our study may explain the lack of systemic inflammation, and differences in STMC populations. Elevation in activated-macrophages is common in ST from late-stage OA-patients,[17, 20, 21] and our cell depletion were consistent with a pro-catabolic role of ST-macrophages. However, the obesity/metabolic-syndrome common in these patients can itself cause inflammation with macrophage accumulation.[49-51] Obesity/metabolic-syndrome does worsen surgically-induced ptOA in mice,[51-54] and pre-operative depletion of ST-macrophages did reduce synovitis and structural pathology post-DMM in obese C57BL6 mice.[51] However, in other studies, despite systemic- and ST-macrophage depletion immediately post-DMM, there was no change in structural disease and actually increased joint inflammation and ST-CD3^+^ T-cell influx.[55] Local macrophage depletion in non-obese animals reduces structural joint pathology in arthritis models with a more profound inflammatory pathophysiology,[56-58] but data in surgically-induced ptOA in non-obese mice is lacking. Both DMM- and Sham-surgery in our non-obese mice led to equivalent ST-monocyte/macrophage infiltration and activation that resolved irrespective of OA onset or progression. This suggests that ST-macrophage accumulation in OA-patients and STR/ort mice may be associated more with their meta-inflammatory co-morbidity than the OA *per se*.[59]

In contrast to macrophages, we showed major differences between DMM and Sham in ST-lymphocytes. Our data in advanced ptOA (day-112) are in accordance with that from end-stage OA-patients showing the presence and activated phenotype of ST-CD4^+^T-cells.[19-22] We now report that increased ST-lymphocytes, particularly CD4^+^, occurs almost immediately following OA-inducing joint injury, consistent with the limited available data in patients with early versus late OA.[9] The number of ST-CD8^+^ T-cells was also increased in DMM compared with NOC and Sham up to 8 weeks post-surgery but there was less difference immediately post-surgery or in late-stage OA. There is some suggestion in patients that ST-CD8^+^T-cells increase with knee OA severity, [60] and we found more patients with late OA had detectable *CD8* mRNA in their ST. There may be confounding effects as both age and obesity/metabolic-syndrome increase CD8^+^T-cell number and activation in blood and peripheral tissues,[61-63] and lack of systemic lymphocyte change or increase in late-stage ST-CD8^+^T-cells in our study may be associated with use of young non-obese mice.

Previous studies have demonstrated a preponderance of Th1-activated ST-CD4^+^T-cells in late-stage human OA, with lower percentages of Treg, Th2, and Th17.[22] Our data shows that ST-Th1/Th2 imbalance occurs particularly during early and progressive OA, and then diminishes before becoming apparent again in late-stage disease. In contrast, ST-Th17/Treg imbalance predominated acutely post-injury, was lost in early disease, and then re-emerged later due to a decrease in Tregs. Increased ST-Th17 has been reported in OA patients,[64] but rather than being decreased, Tregs were found in higher numbers in ST than blood and with more advanced radiographic disease.[22, 65, 66] Again differences in Th17/Treg balance in late-stage OA in DMM versus patients is likely associated with confounding effects of obesity and ageing in the latter.[63]

Lymphocyte cell-surface-marker mRNA expression can provide a surrogate for flow-cytometric cell counting.[10] That expression of classic monocyte/macrophage cytokines *Il1* and *Il6* in ST was not higher in DMM than Sham and returned to naïve levels by 5-weeks post-surgery (Supplementary Figure 1A), was consistent with the lack of OA-specific innate immune-cell response (Figure 2). ST mRNA expression of *Cd4, Cd8*, and the Th1-cytokine *Tnf* (Supplementary Figure 1B) showed cyclic changes post-surgery and were increased in DMM similar to flow-cytometry, supporting use of ST-mRNA for analysis of archival human ST samples. While mean mRNA levels of lymphocyte markers were not different between patient cohorts as in previous analysis,[10] the temporal pattern in the meniscal injury groups was consistent with the mouse data, showing an early CD3^+^CD8^+^ response. Of interest was the positive association between ST *CD8* but not *CD4* expression with pain/symptoms in meniscal injury patients. Generalised synovitis is associated with worse symptoms in patients with established OA,[8] and meniscal injury.[26, 27] In late-stage OA-patients, the percentage of ST-CD4^+^T-cells, but not CD8^+^T-cells or macrophages, has been associated with pain even after adjusting for BMI.[20] While CD4^+^T-cells have been strongly implicated in chronic neuropathic pain pathophysiology, the role of CD8^+^T-cells is less clear with both pro- and anti-algesic effects reported.[67, 68] Our association data in patients suggests a novel role for synovial CD8^+^T-cells in the early clinical symptoms following meniscal injury, but pre- or post-DMM anti-CD3 failed to modify pain-sensitization in mice (Supplementary Figure 2). This could be associated with sex differences as T-cells may play a more important role in chronic pain hypersensitivity in females,[69] or because systemic anti-CD3 failed to modify lymphocyte numbers in the ST where they may play a more direct pro-algesic role. Finally, our broad anti-CD3 depletion strategy may have affected both pro- and anti-algesic T-cells, and a more targeted approach may be needed.

Synovitis increases the risk of incident structural OA in population-based studies,[12, 13] and progression of cartilage damage in patients with meniscal injuries.[70] Our patient ST-cell depletion studies demonstrate that CD3^+^lymphocytes play a role in upregulation of patho-physiologically relevant MMPs, consistent with previous reports.[10] Whether this is through inhibiting synthesis of metalloproteinases by lymphocytes or modulatuion of their co-stimulation of macrophage MMP synthesis is unclear.[71, 72] CD4^+^T-cell and macrophage activation in OA synovial fluid are positively associated,[17] and compared with PBMCs T-cells in late-OA ST show increased activation and secretion of inflammatory cytokines and chemokines.[20, 22, 64, 73, 74] This suggests that factors in the local joint environment drive T-cell cytokine secretion, which in turn may modulate the effects of activated CD4^+^T-cells in increasing chondrocyte MMP expression.[74]

Decreased lymphatic drainage occurs in surgically induced ptOA onset in mice, with synovitis, osteophytes and cartilage damage worsened by inhibition of lymph-angiogenesis and ameliorated with improvement in lymphatic function.[75]. More direct evidence for T-cells in OA pathophysiology comes from mice deficient in CD4[76] or CD8[77] which both have reduced ptOA cartilage damage, and despite constitutive depletion only protection in late-stage ptOA was seen. This is in line with our anti-CD3 data, but by using a transient depletion strategy we show that while the immediate and 4-week post-DMM T-cell response contribute to osteophyte development, the earliest T-cell response is key to late stage ptOA cartilage damage. This is consistent with the presence and severity of the initial synovitis following meniscal injury even if it subsequently resolves, being associated with risk of progressive cartilage damage in patients.[70] While there was no change in PBMC or SpMC T-cell number or activation in DMM-induced ptOA, the anti-CD3 treatment outcomes implicate systemic T-cells in progressive cartilage damage. That systemic T-cell depletion was not mirrored in the ST raises the question of where the ST-cells originate. As well as influx and polarization of circulating cells, activation and proliferation of resident populations could contribute to both the innate- and adaptive-immune response. The majority of CD45^+^ cells in normal ST are CD11b^+^ monocyte-lineage, and our current and previous data[10, 51] indicate that ∼20% of these are macrophages that could participate immediately post-injury. The predominant resident-monocyte population could then be activated by mechanical, cellular or soluble stimuli.[11, 33, 36, 37, 59] As the number of resident T-cells in normal ST is very low, the immediate post-injury increase more likely arises through influx rather than local replication. Analysis of late-stage OA patients indicates CD8^+^ clonality but whether this occurs through selective migration/influx or local clonal selection/expansion is unclear.[64]

In conclusion our comprehensive temporal quantitative comparison of ST inflammatory cell influx after OA-inducing versus non-OA-inducing joint injury, has enabled us to define the important role of the adaptive immune response. Increases in number and activation of ST-T-cells in joints at risk of OA occurred within the first 3 days of injury, and while recurring cyclically through subsequent disease onset, depletion studies indicated this initial influx was key to long-term ptOA risk. There are a number of differences between our pre-clinical animal and patient populations, that highlight key research questions for further study. Thus, the precise local environmental cues in the OA joint that stimulate and maintain the adaptive-immune response, the specific T-cell subsets involved in symptomatic and structural disease, and modification by systemic factors such as age, sex, and obesity/metabolic-syndrome remain to be defined. Nevertheless, the present results identify a hitherto unappreciated pathophysiological role of acute T-cell activation after joint injury in ptOA risk, opening new diagnostic, prognostic and therapeutic avenues.

## Data Availability

All data associated with this study are present in the paper or the Supplementary Materials.

## Acknowledgements

We thank Craig Della Valle, Charles Bush-Joseph and Nikhil Verma (Rush University Medical Center, Department of Orthopedics, Chicago, IL), and Brian McKeon and Anthony Albert (New England Baptist Hospital, Department of Orthopedics, Boston, MA USA) for the collection of the human synovial tissue biopsies.

## Author contributions

Babak Moradi, Miriam Jackson, and Christopher Little designed the study. Experimental data was generated by Babak Moradi, Miriam Jackson, Cindy Shu, Susan Smith, Sanaa Zaki, Hadrian Platzer, Nils Rosshirt, David Giangreco and Carla Scanzello. Statistical analysis performed by Babak Moradi, Miriam Jackson and Margaret Smith. The paper was drafted by Babak Moradi, Miriam Jackson, and Christopher Little, and all authors contributed to editing and approved the final version.

## Data and materials availability

All data associated with this study are present in the paper or the Supplementary Materials.

## Patient and Public Involvement

While data from patients is included in the study (with appropriate institutional ethics approval), patients/consumers were not involved in the design, conduct, reporting, or dissemination plans of our research

## Supplementary Information

### Cohorts for human ST mRNA analysis

All studies were approved by the Institutional Review Boards (IRB) of Rush University Medical Center or the New England Baptist Hospital (NEBH), and all patients gave written, informed consent.

#### Advanced knee OA

16 synovial tissue specimens were obtained from patients with advanced OA undergoing total knee replacement, who had provided tissue specimens to the IRB-approved (Protocol Nr. L00011021) Orthopedic Tissue and Implant Repository at Rush University Medical Center (Chicago, IL). All specimens came from patients meeting ACR criteria for knee OA confirmed by the operating surgeon and had radiographic evidence of moderate to severe knee OA on pre-operative knee x-rays (Kellgren-Lawrence score >2). Age, sex, and BMI were available from these patients in addition to radiographic score.

#### Meniscal injury with early OA (Rush)

19 synovial specimens from patients with early-stage knee OA undergoing arthroscopic partial meniscectomy were obtained from the IRB-approved Knee Injury and Arthritis repository (Protocol Nr. 10011306), also located at Rush University Medical Center (Chicago, IL). The diagnosis of early-stage OA was determined by the operating surgeon, and confirmed by presence of a degenerative meniscal tear and either intraoperative evidence of cartilage degeneration in the medial, lateral or patellofemoral compartments (Outerbridge score ≥1), OR presence of radiographic OA changes (Kellgren-Lawrence stage ≥1) on pre-operative x-rays. Age, gender, BMI, and location of meniscal tear were available for these patients, and the Knee Injury and Osteoarthritis Outcome Score (KOOS), English version LK1.0 was used to measure preoperative knee symptoms. Questionnaires, scoring manual and user’s guide were obtained from http://www.koos.nu. The KOOS measures symptoms and disability on five separately scored sub scales: Pain, other Symptoms, Activities of Daily Living (ADL), Sports and Recreation and Quality of Life (QOL).

#### Meniscal injury with no OA (NEBH)

13 synovial tissue specimens were obtained from patients who had participated in a longitudinal study at the New England Baptist Hospital (Boston, MA). These patients had a history of acute knee injury within the previous 1 year, had an MRI-confirmed meniscal tear and had undergone arthroscopic partial meniscectomy. Patients with any radiographic evidence of OA (Kellgren-Lawrence score >0) were excluded from this study. Age, sex, BMI, location of meniscal tear and pre-operative Lysholm scores were available for these patients. The Lysholm questionnaire is a clinician-developed instrument, measuring symptoms including pain, swelling, limp, locking, and instability, as well as functional disability (stair-climbing, squatting, use of supports). A summed score is reported on a scale of 0-100, where 100 = no symptoms/disability.

**Supplementary Figure 1: Changes in synovial cytokine expression following sham or DMM surgery, and effects of T-cell depletion on pain. (A)** Expression of *Il1* and *Il6* mRNA in mouse synovial tissue was markedly increased 7-days post-DMM and then declined toward NOC levels by day 35-49. While expression of both cytokines was similar in Sham and DMM at day-7, it remained elevated in Sham at day-14 (>DMM) before declining. **(B)** While *Tnf* expression also increased at day 7 and declined at day 14 in Sham and DMM, it showed a second peak elevation at day-49 and remained higher than NOC in both operated joints (DMM > Sham) for the remainder of the study. **(C)** Peripheral tactile allodynia measured as significant reduction in 50% withdrawal reflex using with Von Frey fibres, was evident 2 weeks post-DMM and remained present through the course of the study. There was no difference in allodynia at any time between mice treated Pre- or Post-DMM with IgG versus anti-CD3. Data is shown as the mean ± SEM; NOC = triangle and solid line, Sham = circle and dotted line, DMM = square and dashed line. Statistical comparison was performed for those time points where scores for all groups were available (highlighted with a vertical dashed line); n = 5-6 individual mouse synovial tissues/time/treatment, and repeated measure in n = 8 mice/treatment for allodynia; *=*P*≤0.05; +=*P*≤0.01; #=*P*≤0.001.

**Supplementary Table I.**
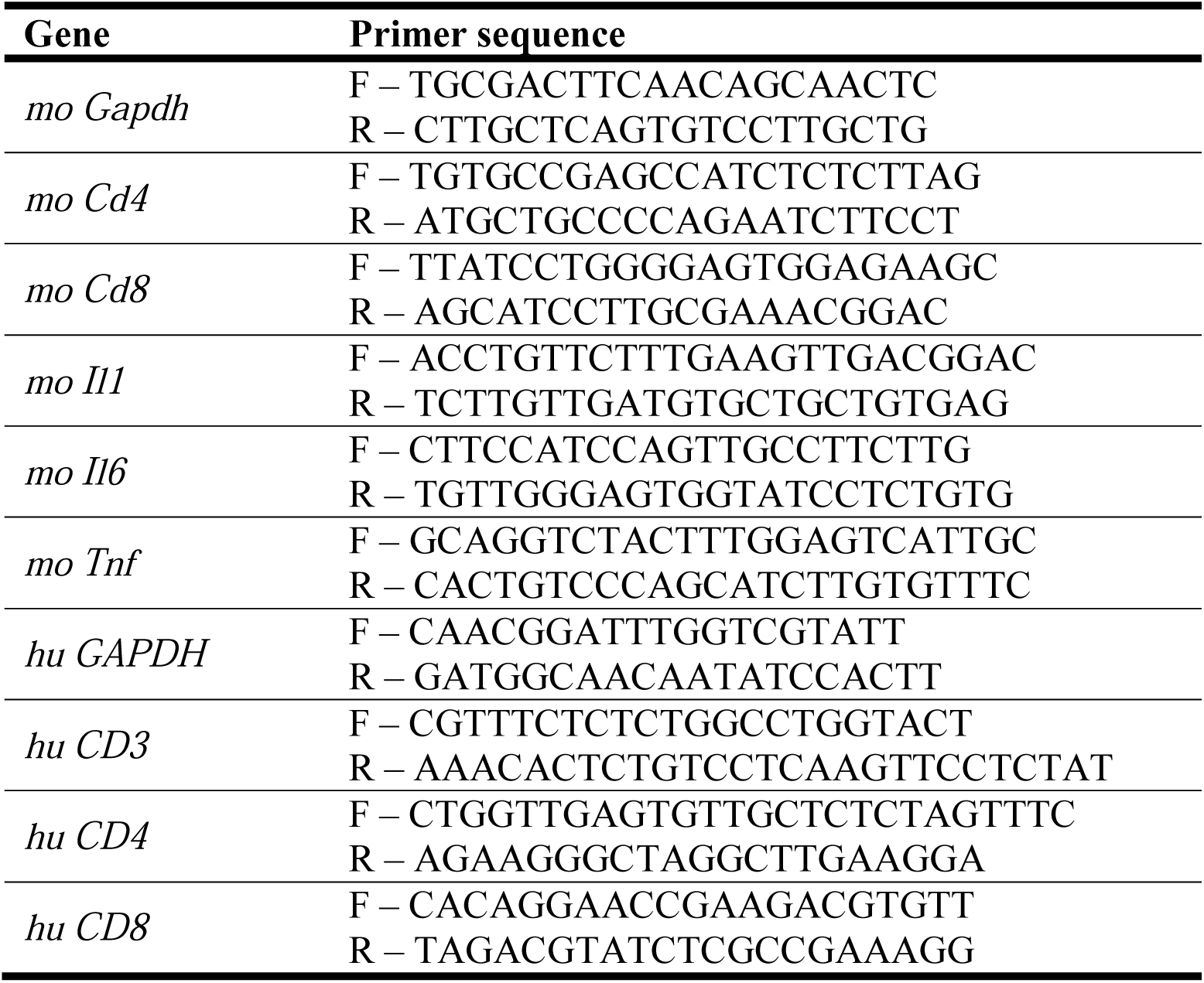
Mouse (mo) or human (hu) genes with their specific forward (F) and reverse (R) PCR primer sequences.

**Supplementary Table II:**
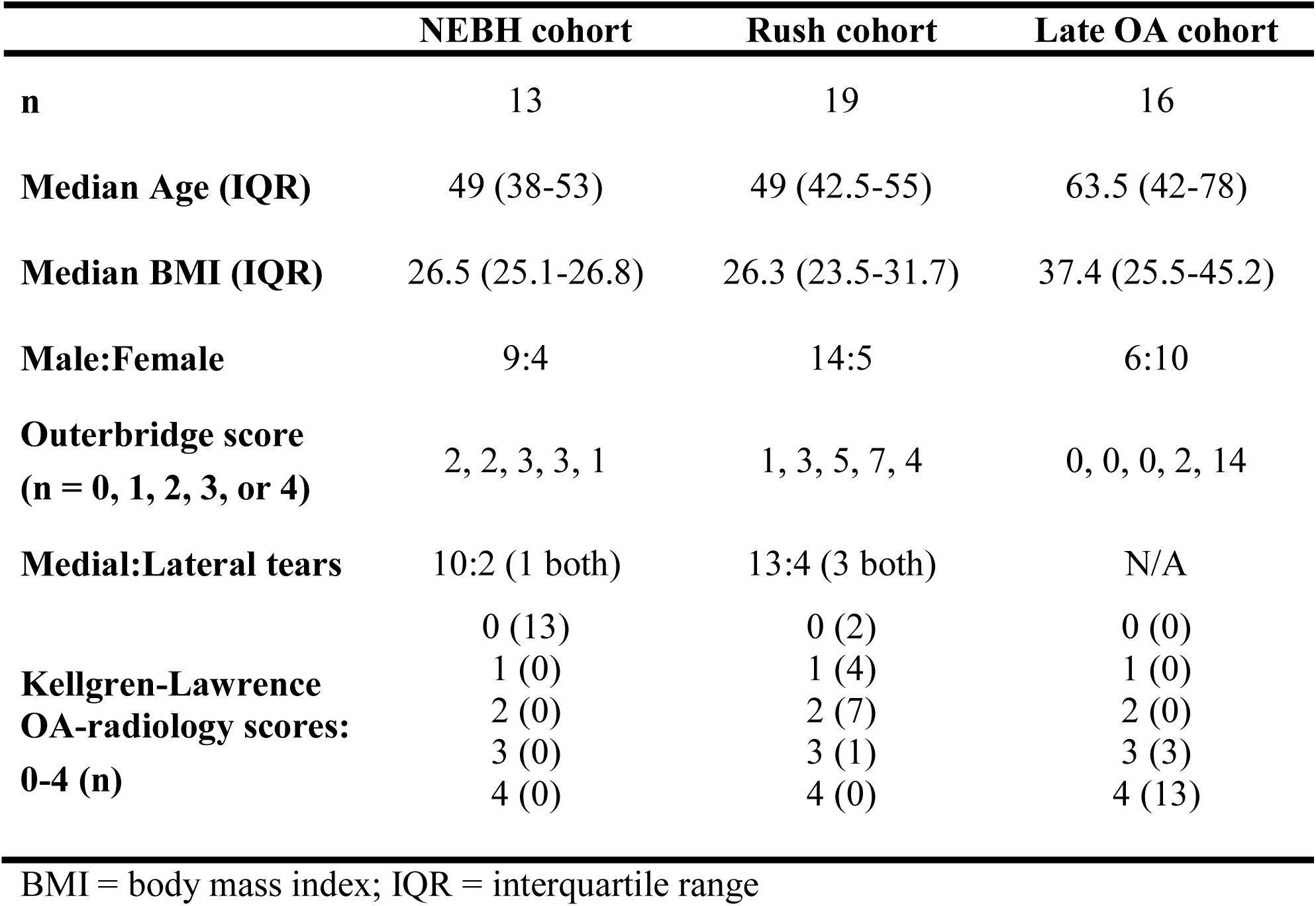
Characteristics of meniscal injury and OA patient cohorts used for synovial tissue mRNA analysis.

**Supplementary Table III:** Characteristics of OA patients whose synovial tissue was used for cell depletion studies.

|  |  |
| --- | --- |
| **Number of patients** | 12 |
| **Gender, n (%)** |  |
| Male | 3 (25 %) |
| Female | 9 (75 %) |
| **Age at surgery (years), Mean ± SD** | 69.0 ± 12.6 |
| **Knee side, n (%)** |  |
| Right | 6 (50 %) |
| Left | 6 (50 %) |
| **Operation type** |  |
| Uni-compartmental knee arthroplasty, n (%) | 7 (58.3%) |
| Total knee arthroplasty, n (%) | 5 (41.7 %) |
| **BMI (kg/m<sup>2</sup>), mean ± SD** | 26.0 ± 3.2 |
| **Synovial leucocytes cells/nl, mean ± SD** | 7.2 ± 2.2 |
| **Serum C-reactive protein mg/l, mean ± SD** | 2.4 ± 0.87 |
| **Kellgren-Lawrence OA-radiology score, n (%)** |  |
| 2 | 1 (8.3 %) |
| 3 | 3 (25 %) |
| 4 | 8 (66.7 %) |

BMI = body mass index; IQR = interquartile range

**Supplementary Table IV:** Comparison of the ratios of Th1/Th2 and Th17/Treg CD4 T-cells in the spleen versus synovium of mice at different times (day 7, 35, 56, 70, 84, 98 and 112) following DMM surgery. Data shows the ratios calculated from the mean cell number data in Table I. Figures highlighted in bold text are those where significant differences in cell % were seen in that specific tissue and post-DMM time point (Figure 4B).

| **Time post-DMM** | **Spleen**<br>Th1/Th2 ratio | **Synovium**<br>Th1/Th2 ratio | **Spleen**<br>Th17/Treg ratio | **Synovium**<br>Th17/Treg ratio |
| --- | --- | --- | --- | --- |
| Day 7 | **0.8** | 1.2 | 1.3 | **13.9** |
| Day 35 | 1.4 | **3.3** | **1.9** | 0.8 |
| Day 56 | **0.2** | **43.3** | 0.4 | 0.7 |
| Day 70 | **1.9** | **2.2** | 2.2 | **1.9** |
| Day 84 | **1.5** | 1.4 | **4.0** | **3405.1** |
| Day 98 | **0.5** | **3.3** | 1.1 | **17.4** |
| Day 112 | 1.4 | 8.3 | 1.2 | 2.3 |

**Supplementary Table V:** Spearman correlation (R (*P*-value)) of synovial lymphocyte marker gene expression and pre-operative KOOS clinical scores in patients with meniscal injuries from the Rush cohort, and pre- and post-surgical Lysholm clinical scores for the NEBH cohort. Data in bold typeface is statistically significant.

| Gene | KOOS-Pain<br>(n = 16) | KOOS-Symptoms<br>(n = 17) | KOOS-ADL<br>(n = 16) | KOOS-Sport/Rec<br>(n = 17) | KOOS-QoL<br>(n = 16) |
| --- | --- | --- | --- | --- | --- |
| *CD3* | -0.36 (0.17) | -0.41 (0.10) | -0.37 (0.16) | -0.35 (0.16) | -0.48 (0.06) |
| *CD4* | 0.07 (0.81) | -0.04 (0.84) | 0.11 (0.67) | 0.01 (0.98) | 0.28 (0.29) |
| *CD8* | **-0.47 (0.05)** | -.033 (0.13) | **-0.65 (0.004)** | **-0.57 (0.01)** | **-0.57 (0.01)** |
|  | Pre-surgery Lysholm<br>(n =13) | 16 wk post-surgery<br>Lysholm (n =12) | 1 yr post-surgery<br>Lysholm (n = 9) | 2 yr post-surgery<br>Lysholm (n =13) |  |
| *CD3* | **-0.64 (0.02)** | -0.02 (0.93) | -0.45 (0.22) | 0.09 (0.77) |  |
| *CD4* | -0.31 (0.30) | -0.25 (0.41) | -0.23 (0.54) | 0.09 (0.77) |  |
| *CD8* | -0.44 (0.13) | 0.16 (0.61) | -0.37 (0.31) | 0.10 (0.76) |  |

## References

1. Ackerman IN, Pratt C, Gorelik A, Liew D. Projected Burden of Osteoarthritis and Rheumatoid Arthritis in Australia: A Population-Level Analysis. Arthritis Care Res (Hoboken). 2018 Jun; 70(6):877–883.

2. Menon J, Mishra P. Health care resource use, health care expenditures and absenteeism costs associated with osteoarthritis in US healthcare system. Osteoarthritis Cartilage. 2018 Apr; 26(4):480–484.

3. de Lange-Brokaar BJ, Ioan-Facsinay A, van Osch GJ, Zuurmond AM, Schoones J, Toes RE, et al. Synovial inflammation, immune cells and their cytokines in osteoarthritis: a review. Osteoarthritis Cartilage. 2012 Dec; 20(12):1484–1499.

4. Livshits G, Kalinkovich A. Hierarchical, imbalanced pro-inflammatory cytokine networks govern the pathogenesis of chronic arthropathies. Osteoarthritis Cartilage. 2018 Jan; 26(1):7–17.

5. Lopes EBP, Filiberti A, Husain SA, Humphrey MB. Immune Contributions to Osteoarthritis. Curr Osteoporos Rep. 2017 Dec; 15(6):593–600.

6. Mathiessen A, Conaghan PG. Synovitis in osteoarthritis: current understanding with therapeutic implications. Arthritis Res Ther. 2017 Feb 2; 19(1):18.

7. Scanzello CR. Role of low-grade inflammation in osteoarthritis. Curr Opin Rheumatol. 2017 Jan; 29(1):79–85.

8. Wang X, Hunter DJ, Jin X, Ding C. The importance of synovial inflammation in osteoarthritis: current evidence from imaging assessments and clinical trials. Osteoarthritis Cartilage. 2018 Feb; 26(2):165–174.

9. Benito MJ, Veale DJ, FitzGerald O, van den Berg WB, Bresnihan B. Synovial tissue inflammation in early and late osteoarthritis. Ann Rheum Dis. 2005 Sep; 64(9):1263–1267.

10. Scanzello CR, Umoh E, Pessler F, Diaz-Torne C, Miles T, Dicarlo E, et al. Local cytokine profiles in knee osteoarthritis: elevated synovial fluid interleukin-15 differentiates early from end-stage disease. Osteoarthritis Cartilage. 2009 Aug; 17(8):1040–1048.

11. Wang Q, Rozelle AL, Lepus CM, Scanzello CR, Song JJ, Larsen DM, et al. Identification of a central role for complement in osteoarthritis. Nat Med. 2011 Dec; 17(12):1674–1679.

12. Atukorala I, Kwoh CK, Guermazi A, Roemer FW, Boudreau RM, Hannon MJ, et al. Synovitis in knee osteoarthritis: a precursor of disease? Ann Rheum Dis. 2016 Feb; 75(2):390–395.

13. Roemer FW, Guermazi A, Felson DT, Niu J, Nevitt MC, Crema MD, et al. Presence of MRI-detected joint effusion and synovitis increases the risk of cartilage loss in knees without osteoarthritis at 30-month follow-up: the MOST study. Ann Rheum Dis. 2011 Oct; 70(10):1804–1809.

14. Davis JE, Ward RJ, MacKay JW, Lu B, Price LL, McAlindon TE, et al. Effusion-synovitis and infrapatellar fat pad signal intensity alteration differentiate accelerated knee osteoarthritis. Rheumatology (Oxford). 2019 Mar 1; 58(3):418–426.

15. Lust G, Summers BA. Early, asymptomatic stage of degenerative joint disease in canine hip joints. Am J Vet Res. 1981 Nov; 42(11):1849–1855.

16. Little CB, Hunter DJ. Post-traumatic osteoarthritis: from mouse models to clinical trials. Nat Rev Rheumatol. 2013; 9(8):485–497.

17. Gomez-Aristizabal A, Gandhi R, Mahomed NN, Marshall KW, Viswanathan S. Synovial fluid monocyte/macrophage subsets and their correlation to patient-reported outcomes in osteoarthritic patients: a cohort study. Arthritis Res Ther. 2019 Jan 18; 21(1):26.

18. Kraus VB, McDaniel G, Huebner JL, Stabler TV, Pieper CF, Shipes SW, et al. Direct in vivo evidence of activated macrophages in human osteoarthritis. Osteoarthritis Cartilage. 2016 Sep; 24(9):1613–1621.

19. Ishii H, Tanaka H, Katoh K, Nakamura H, Nagashima M, Yoshino S. Characterization of infiltrating T cells and Th1/Th2-type cytokines in the synovium of patients with osteoarthritis. Osteoarthritis Cartilage. 2002 Apr; 10(4):277–281.

20. Klein-Wieringa IR, de Lange-Brokaar BJ, Yusuf E, Andersen SN, Kwekkeboom JC, Kroon HM, et al. Inflammatory Cells in Patients with Endstage Knee Osteoarthritis: A Comparison between the Synovium and the Infrapatellar Fat Pad. J Rheumatol. 2016 Apr; 43(4):771–778.

21. Moradi B, Rosshirt N, Tripel E, Kirsch J, Barie A, Zeifang F, et al. Unicompartmental and bicompartmental knee osteoarthritis show different patterns of mononuclear cell infiltration and cytokine release in the affected joints. Clin Exp Immunol. 2015 Apr; 180(1):143–154.

22. Rosshirt N, Hagmann S, Tripel E, Gotterbarm T, Kirsch J, Zeifang F, et al. A predominant Th1 polarization is present in synovial fluid of end-stage osteoarthritic knee joints: analysis of peripheral blood, synovial fluid and synovial membrane. Clin Exp Immunol. 2019 Mar; 195(3):395–406.

23. Lieberthal J, Sambamurthy N, Scanzello CR. Inflammation in joint injury and post-traumatic osteoarthritis. Osteoarthritis Cartilage. 2015 Nov; 23(11):1825–1834.

24. Nair A, Gan J, Bush-Joseph C, Verma N, Tetreault MW, Saha K, et al. Synovial chemokine expression and relationship with knee symptoms in patients with meniscal tears. Osteoarthritis Cartilage. 2015 Jul; 23(7):1158–1164.

25. Pessler F, Dai L, Diaz-Torne C, Gomez-Vaquero C, Paessler ME, Zheng DH, et al. The synovitis of “non-inflammatory” orthopaedic arthropathies: a quantitative histological and immunohistochemical analysis. Ann Rheum Dis. 2008 Aug; 67(8):1184–1187.

26. Scanzello CR, Albert AS, DiCarlo E, Rajan KB, Kanda V, Asomugha EU, et al. The influence of synovial inflammation and hyperplasia on symptomatic outcomes up to 2 years post-operatively in patients undergoing partial meniscectomy. Osteoarthritis Cartilage. 2013 Sep; 21(9):1392–1399.

27. Scanzello CR, McKeon B, Swaim BH, Dicarlo E, Asomugha EU, Kanda V, et al. Synovial inflammation in patients undergoing arthroscopic meniscectomy: Molecular characterization and relationship to symptoms. Arthritis Rheum. 2011 Feb; 63(2):391–400.

28. Watt FE, Paterson E, Freidin A, Kenny M, Judge A, Saklatvala J, et al. Acute Molecular Changes in Synovial Fluid Following Human Knee Injury: Association With Early Clinical Outcomes. Arthritis & rheumatology (Hoboken, NJ). 2016 Sep; 68(9):2129–2140.

29. Malfait AM, Little CB. On the predictive utility of animal models of osteoarthritis. Arthritis Res Ther. 2015; 17:225.

30. Jackson MT, Moradi B, Zaki S, Smith MM, McCracken S, Smith SM, et al. Depletion of protease-activated receptor 2 but not protease-activated receptor 1 may confer protection against osteoarthritis in mice through extracartilaginous mechanisms. Arthritis & rheumatology (Hoboken, NJ). 2014 Dec; 66(12):3337–3348.

31. Shu CC, Jackson MT, Smith MM, Smith SM, Penm S, Lord MS, et al. Ablation of Perlecan Domain 1 Heparan Sulfate Reduces Progressive Cartilage Degradation, Synovitis, and Osteophyte Size in a Preclinical Model of Posttraumatic Osteoarthritis. Arthritis & rheumatology (Hoboken, NJ). 2016 Apr; 68(4):868–879.

32. Chaplan SR, Bach FW, Pogrel JW, Chung JM, Yaksh TL. Quantitative assessment of tactile allodynia in the rat paw. J Neurosci Methods. 1994 Jul; 53(1):55–63.

33. Nair A, Kanda V, Bush-Joseph C, Verma N, Chubinskaya S, Mikecz K, et al. Synovial fluid from patients with early osteoarthritis modulates fibroblast-like synoviocyte responses to toll-like receptor 4 and toll-like receptor 2 ligands via soluble CD14. Arthritis Rheum. 2012 Jul; 64(7):2268–2277.

34. Kung LHW, Ravi V, Rowley L, Bell KM, Little CB, Bateman JF. Comprehensive Expression Analysis of microRNAs and mRNAs in Synovial Tissue from a Mouse Model of Early Post-Traumatic Osteoarthritis. Scientific reports. 2017 Dec 18; 7(1):17701.

35. Smolen JS, Breedveld FC, Burmester GR, Bykerk V, Dougados M, Emery P, et al. Treating rheumatoid arthritis to target: 2014 update of the recommendations of an international task force. Ann Rheum Dis. 2016 Jan; 75(1):3–15.

36. van den Bosch MHJ. Inflammation in osteoarthritis: is it time to dampen the alarm(in) in this debilitating disease? Clin Exp Immunol. 2019 Feb; 195(2):153–166.

37. Scanzello CR, Plaas A, Crow MK. Innate immune system activation in osteoarthritis: is osteoarthritis a chronic wound? Curr Opin Rheumatol. 2008 Sep; 20(5):565–572.

38. Larouche J, Sheoran S, Maruyama K, Martino MM. Immune Regulation of Skin Wound Healing: Mechanisms and Novel Therapeutic Targets. Advances in wound care. 2018 Jul 1; 7(7):209–231.

39. Li YS, Luo W, Zhu SA, Lei GH. T Cells in Osteoarthritis: Alterations and Beyond. Frontiers in immunology. 2017; 8:356.

40. de Jong H, Berlo SE, Hombrink P, Otten HG, van Eden W, Lafeber FP, et al. Cartilage proteoglycan aggrecan epitopes induce proinflammatory autoreactive T-cell responses in rheumatoid arthritis and osteoarthritis. Ann Rheum Dis. 2010 Jan; 69(1):255–262.

41. Brophy RH, Tycksen ED, Sandell LJ, Rai MF. Changes in Transcriptome-Wide Gene Expression of Anterior Cruciate Ligament Tears Based on Time From Injury. Am J Sports Med. 2016 Aug; 44(8):2064–2075.

42. Burleigh A, Chanalaris A, Gardiner MD, Driscoll C, Boruc O, Saklatvala J, et al. Joint immobilization prevents murine osteoarthritis and reveals the highly mechanosensitive nature of protease expression in vivo. Arthritis Rheum. 2012 Jul; 64(7):2278–2288.

43. Lefevre S, Knedla A, Tennie C, Kampmann A, Wunrau C, Dinser R, et al. Synovial fibroblasts spread rheumatoid arthritis to unaffected joints. Nat Med. 2009 Dec; 15(12):1414–1420.

44. Staines KA, Poulet B, Wentworth DN, Pitsillides AA. The STR/ort mouse model of spontaneous osteoarthritis - an update. Osteoarthritis Cartilage. 2017 Jun; 25(6):802–808.

45. Uchida K, Naruse K, Satoh M, Onuma K, Ueno M, Takano S, et al. Increase of circulating CD11b(+)Gr1(+) cells and recruitment into the synovium in osteoarthritic mice with hyperlipidemia. Exp Anim. 2013; 62(3):255–265.

46. Kyostio-Moore S, Nambiar B, Hutto E, Ewing PJ, Piraino S, Berthelette P, et al. STR/ort mice, a model for spontaneous osteoarthritis, exhibit elevated levels of both local and systemic inflammatory markers. Comp Med. 2011 Aug; 61(4):346–355.

47. Uchida K, Urabe K, Naruse K, Ogawa Z, Mabuchi K, Itoman M. Hyperlipidemia and hyperinsulinemia in the spontaneous osteoarthritis mouse model, STR/Ort. Exp Anim. 2009 Apr; 58(2):181–187.

48. Berenbaum F, Wallace IJ, Lieberman DE, Felson DT. Modern-day environmental factors in the pathogenesis of osteoarthritis. Nat Rev Rheumatol. 2018 Nov; 14(11):674–681.

49. Barboza E, Hudson J, Chang WP, Kovats S, Towner RA, Silasi-Mansat R, et al. Profibrotic Infrapatellar Fat Pad Remodeling Without M1 Macrophage Polarization Precedes Knee Osteoarthritis in Mice With Diet-Induced Obesity. Arthritis & rheumatology (Hoboken, NJ). 2017 Jun; 69(6):1221–1232.

50. Hamada D, Maynard R, Schott E, Drinkwater CJ, Ketz JP, Kates SL, et al. Suppressive Effects of Insulin on Tumor Necrosis Factor-Dependent Early Osteoarthritic Changes Associated With Obesity and Type 2 Diabetes Mellitus. Arthritis & rheumatology (Hoboken, NJ). 2016 Jun; 68(6):1392–1402.

51. Sun AR, Wu X, Liu B, Chen Y, Armitage CW, Kollipara A, et al. Pro-resolving lipid mediator ameliorates obesity induced osteoarthritis by regulating synovial macrophage polarisation. Scientific reports. 2019 Jan 23; 9(1):426.

52. Guss JD, Ziemian SN, Luna M, Sandoval TN, Holyoak DT, Guisado GG, et al. The effects of metabolic syndrome, obesity, and the gut microbiome on load-induced osteoarthritis. Osteoarthritis Cartilage. 2019 Jan; 27(1):129–139.

53. Louer CR, Furman BD, Huebner JL, Kraus VB, Olson SA, Guilak F. Diet-induced obesity significantly increases the severity of posttraumatic arthritis in mice. Arthritis Rheum. 2012 Oct; 64(10):3220–3230.

54. Mooney RA, Sampson ER, Lerea J, Rosier RN, Zuscik MJ. High-fat diet accelerates progression of osteoarthritis after meniscal/ligamentous injury. Arthritis Res Ther. 2011; 13(6):R198.

55. Wu CL, McNeill J, Goon K, Little D, Kimmerling K, Huebner J, et al. Conditional Macrophage Depletion Increases Inflammation and Does Not Inhibit the Development of Osteoarthritis in Obese Macrophage Fas-Induced Apoptosis-Transgenic Mice. Arthritis & rheumatology (Hoboken, NJ). 2017 Sep; 69(9):1772–1783.

56. Richards PJ, Williams AS, Goodfellow RM, Williams BD. Liposomal clodronate eliminates synovial macrophages, reduces inflammation and ameliorates joint destruction in antigen-induced arthritis. Rheumatology (Oxford). 1999 Sep; 38(9):818–825.

57. Van Lent PL, Holthuysen AE, Van Rooijen N, Van De Putte LB, Van Den Berg WB. Local removal of phagocytic synovial lining cells by clodronate-liposomes decreases cartilage destruction during collagen type II arthritis. Ann Rheum Dis. 1998 Jul; 57(7):408–413.

58. Blom AB, van Lent PL, Libregts S, Holthuysen AE, van der Kraan PM, van Rooijen N, et al. Crucial role of macrophages in matrix metalloproteinase-mediated cartilage destruction during experimental osteoarthritis: involvement of matrix metalloproteinase 3. Arthritis Rheum. 2007 Jan; 56(1):147–157.

59. Li C, Xu MM, Wang K, Adler AJ, Vella AT, Zhou B. Macrophage polarization and meta-inflammation.Translational research : the journal of laboratory and clinical medicine. 2018 Jan; 191:29–44.

60. Apinun J, Sengprasert P, Yuktanandana P, Ngarmukos S, Tanavalee A, Reantragoon R. Immune Mediators in Osteoarthritis: Infrapatellar Fat Pad-Infiltrating CD8+ T Cells Are Increased in Osteoarthritic Patients with Higher Clinical Radiographic Grading. International journal of rheumatology. 2016; 2016:9525724.

61. Nishimura S, Manabe I, Nagasaki M, Eto K, Yamashita H, Ohsugi M, et al. CD8+ effector T cells contribute to macrophage recruitment and adipose tissue inflammation in obesity. Nat Med. 2009 Aug; 15(8):914–920.

62. Ponchel F, Burska AN, Hensor EM, Raja R, Campbell M, Emery P, et al. Changes in peripheral blood immune cell composition in osteoarthritis. Osteoarthritis Cartilage. 2015 Nov; 23(11):1870–1878.

63. Kalathookunnel Antony A, Lian Z, Wu H. T Cells in Adipose Tissue in Aging. Frontiers in immunology. 2018; 9:2945.

64. Sae-Jung T, Sengprasert P, Apinun J, Ngarmukos S, Yuktanandana P, Tanavalee A, et al. Functional and T Cell Receptor Repertoire Analyses of Peripheral Blood and Infrapatellar Fat Pad T Cells in Knee Osteoarthritis. J Rheumatol. 2019 Mar; 46(3):309–317.

65. Moradi B, Schnatzer P, Hagmann S, Rosshirt N, Gotterbarm T, Kretzer JP, et al. CD4(+)CD25(+)/highCD127low/(-) regulatory T cells are enriched in rheumatoid arthritis and osteoarthritis joints--analysis of frequency and phenotype in synovial membrane, synovial fluid and peripheral blood.Arthritis Res Ther. 2014 Apr 17; 16(2):R97.

66. Kriegova E, Manukyan G, Mikulkova Z, Gabcova G, Kudelka M, Gajdos P, et al. Gender-related differences observed among immune cells in synovial fluid in knee osteoarthritis. Osteoarthritis Cartilage. 2018 Sep; 26(9):1247–1256.

67. Baddack-Werncke U, Busch-Dienstfertig M, Gonzalez-Rodriguez S, Maddila SC, Grobe J, Lipp M, et al. Cytotoxic T cells modulate inflammation and endogenous opioid analgesia in chronic arthritis. J Neuroinflammation. 2017 Feb 6; 14(1):30.

68. Yang M, Peyret C, Shi XQ, Siron N, Jang JH, Wu S, et al. Evidence from Human and Animal Studies: Pathological Roles of CD8(+) T Cells in Autoimmune Peripheral Neuropathies. Frontiers in immunology. 2015; 6:532.

69. Sorge RE, Mapplebeck JC, Rosen S, Beggs S, Taves S, Alexander JK, et al. Different immune cells mediate mechanical pain hypersensitivity in male and female mice. Nat Neurosci. 2015 Aug; 18(8):1081–1083.

70. MacFarlane LA, Yang H, Collins JE, Jarraya M, Guermazi A, Mandl LA, et al. Association of Changes in Effusion-Synovitis With Progression of Cartilage Damage Over Eighteen Months in Patients With Osteoarthritis and Meniscal Tear. Arthritis & rheumatology (Hoboken, NJ). 2019 Jan; 71(1):73–81.

71. Bar-Or A, Nuttall RK, Duddy M, Alter A, Kim HJ, Ifergan I, et al. Analyses of all matrix metalloproteinase members in leukocytes emphasize monocytes as major inflammatory mediators in multiple sclerosis. Brain. 2003 Dec; 126(Pt 12):2738–2749.

72. Clark RT, Nance JP, Noor S, Wilson EH. T-cell production of matrix metalloproteinases and inhibition of parasite clearance by TIMP-1 during chronic Toxoplasma infection in the brain. ASN neuro. 2011 Jan 21; 3(1):e00049.

73. Klein-Wieringa IR, Kloppenburg M, Bastiaansen-Jenniskens YM, Yusuf E, Kwekkeboom JC, El-Bannoudi H, et al. The infrapatellar fat pad of patients with osteoarthritis has an inflammatory phenotype. Ann Rheum Dis. 2011 May; 70(5):851–857.

74. Scotece M, Perez T, Conde J, Abella V, Lopez V, Pino J, et al. Adipokines induce pro-inflammatory factors in activated Cd4+ T cells from osteoarthritis patient. J Orthop Res. 2017 Jun; 35(6):1299–1303.

75. Wang W, Lin X, Xu H, Sun W, Bouta EM, Zuscik MJ, et al. Attenuated Joint Tissue Damage Associated With Improved Synovial Lymphatic Function Following Treatment With Bortezomib in a Mouse Model of Experimental Posttraumatic Osteoarthritis. Arthritis & rheumatology (Hoboken, NJ). 2019 Feb; 71(2):244–257.

76. Shen PC, Wu CL, Jou IM, Lee CH, Juan HY, Lee PJ, et al. T helper cells promote disease progression of osteoarthritis by inducing macrophage inflammatory protein-1gamma. Osteoarthritis Cartilage. 2011 Jun; 19(6):728–736.

77. Hsieh JL, Shiau AL, Lee CH, Yang SJ, Lee BO, Jou IM, et al. CD8+ T cell-induced expression of tissue inhibitor of metalloproteinses-1 exacerbated osteoarthritis. Int J Mol Sci. 2013; 14(10):19951–19970.

